# Using body temperature and variables commonly available in the EHR to predict acute infection: A proof-of-concept study showing improved pretest probability estimates for acute COVID-19 infection among discharged emergency department patients

**DOI:** 10.1101/2021.01.21.21250261

**Authors:** Carl T Berdahl, An T Nguyen, Marcio A Diniz, Andrew J Henreid, Teryl K Nuckols, Christopher P Libby, Joshua M Pevnick

## Abstract

**Objectives:** Obtaining body temperature is a quick and easy method to screen for acute infection such as COVID-19. Currently, the predictive value of body temperature for acute infection is inhibited by failure to account for other readily available variables that affect temperature values. In this proof-of-concept study, we sought to improve COVID-19 pretest probability estimation by incorporating covariates known to be associated with body temperature, including patient age, sex, comorbidities, month, time of day.

**Methods:** For patients discharged from an academic hospital emergency department after testing for COVID-19 in March and April of 2020, we abstracted clinical data. We reviewed physician documentation to retrospectively generate estimates of pretest probability for COVID-19. Using patients’ COVID-19 PCR test results as a gold standard, we compared AUCs of logistic regression models predicting COVID-19 positivity that used: 1) body temperature alone; 2) body temperature and pretest probability; 3) body temperature, pretest probability, and body temperature-relevant covariates. Calibration plots and bootstrap validation were used to assess predictive performance for model #3.

**Results:** Data from 117 patients were included. The models’ AUCs were: 1) 0.69 2) 0.72, and 3) 0.76, respectively. The absolute difference in AUC was 0.029 (95%CI −0.057 to 0.114, p=0.25) between model 2 and 1 and 0.038 (95%CI −0.021 to 0.097, p=0.10) between model 3 and 2.

**Conclusions:** By incorporating covariates known to affect body temperature, we demonstrated improved pretest probability estimates of acute COVID-19 infection. Future work should be undertaken to further develop and validate our model in a larger, multi-institutional sample.

## INTRODUCTION

Improving the detection of acute COVID-19 infection is critical to minimizing spread of the infection and limiting avoidable morbidity and mortality.[1] However, the ability to diagnose COVID-19 is often limited because of asymptomatic or mildly symptomatic infection;[2] limited testing availability;[3] and slow turnaround time for test results. Given these limitations, improving pretest probability estimation is a crucial way to improve decision-making about when testing is warranted and thus curb community spread of acute infection.[4]

Measuring body temperature is a quick, easy, and almost ubiquitously available way to evaluate for acute infection. However, the use of temperature to diagnose COVID-19 has not reached its fullest potential for three reasons. First, clinicians tend to use temperature as a binary variable, often with a cutoff of 100.4 degrees Fahrenheit signaling active infection.[5] Thus, many infections can be missed when patients exhibit temperatures that are mildly elevated.[6] Second, failure to account for the effects of other variables on temperature values lead to missed opportunities to detect active infection. Differences in measured body temperature and/or a patient’s ability to mount an elevated temperature in the setting of acute infection occur with: patient age, sex, baseline body temperature, and comorbidities (e.g. depression, allergic conditions); time of day; ambient temperature; calendar month; and route of body temperature measurement.[7-9] Third, our clinical experience suggests that, while clinicians are aware of some of these relationships that should alter the interpretation of temperature values, they probably do not account for them effectively when making clinical decisions.

We propose that it may be possible to improve pretest probability estimation for COVID-19 by developing a novel model that includes variables readily available in the electronic health record (EHR). Figure 1 depicts a conceptual model demonstrating how body temperature-related covariates influence pretest probability, including how a clinician should decide to test for COVID-19 and act on results. Estimating pretest probability is critical to determining which patients should be tested. Additionally, because false-negative rates for COVID-19 testing have been found to range between 2% and 29%,[10] estimating pretest probability has been strongly encouraged to determine which patients should get a second test, even after an initial negative test.[11]

**Figure 1.**
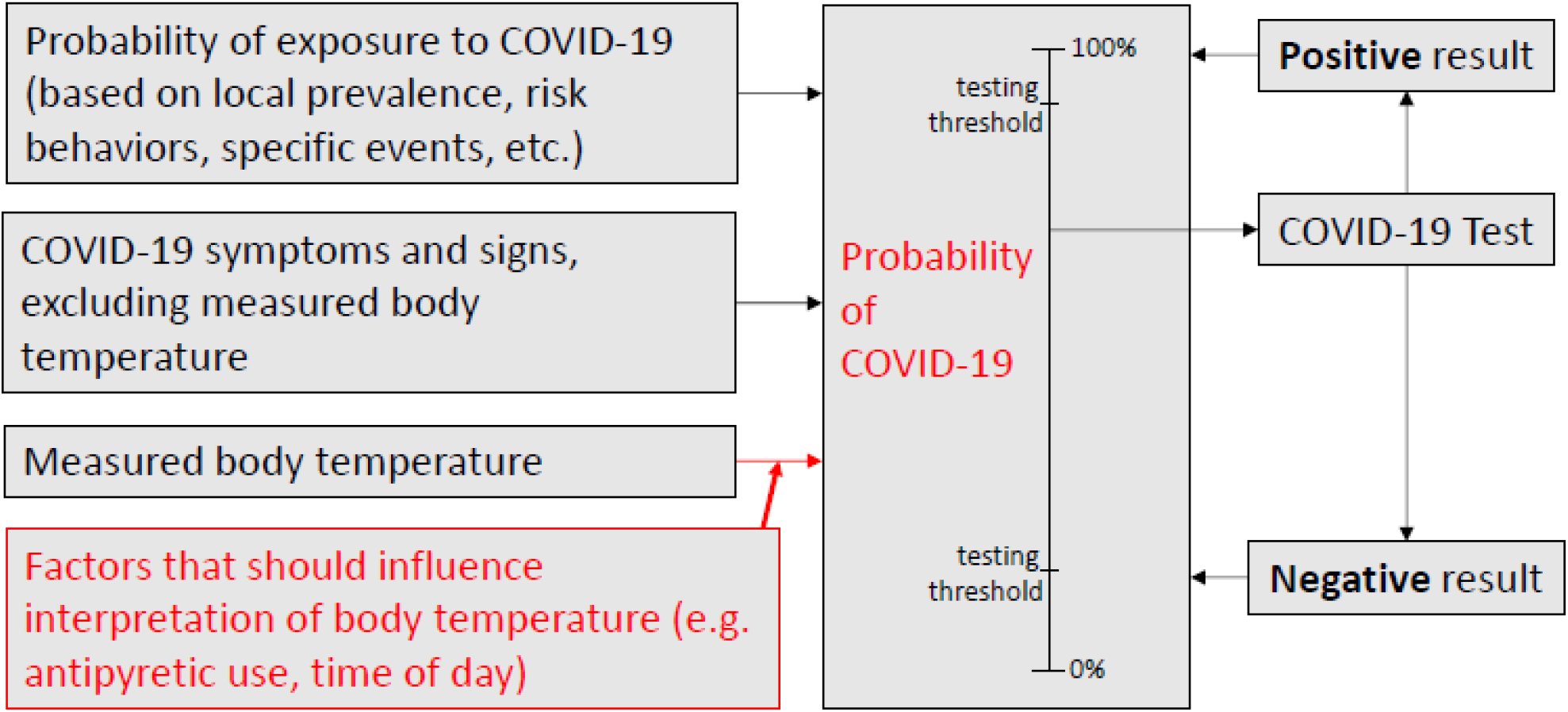
Conceptual model demonstrating how relevant factors (e.g. antipyretic use, time of day) should influence interpretation of body temperature and the resultant effect on pretest probability. Direct influence is shown in red. Note also that in the example shown with a high pretest probability, a positive test result nearly always implies a posttest probability that exceeds the threshold requiring further testing, whereas a negative result may still result in a high enough posttest probability that further testing is warranted.

In this proof-of-concept study of model development, we sought to evaluate whether pretest probability estimates of COVID-19 could be improved by incorporating covariates known to be associated with body temperature, including demographics, comorbidities, time of day, and month of the year. We hypothesized that adding body temperature-relevant covariates would improve the area under the curve (AUC) for a logistic regression model predicting COVID-19 polymerase chain reaction (PCR) test positivity.

## MATERIALS, SUBJECTS, AND METHODS

### Setting and Population

This study was conducted at the emergency department (ED) of an academic medical center in Los Angeles, CA. The study population included patients who were triaged to a temporary COVID-19 surge tent (open from 7:00am to 11:00pm), tested for acute COVID-19 infection with a nasopharyngeal PCR test, and discharged home for outpatient management between March 12 and April 6, 2020. Patients were excluded from the study if they did not have an oral temperature reading, if they were previously seen in the ED within the prior 7 days, if they did not complete ED evaluation (i.e., left without being seen, left against medical advice, or eloped), or if they reported no symptoms.

### Data Sources

Study procedures involved accessing an existing operational database to collect variables characterizing: patient age, sex, vital signs (including date, time, and route of measurement), comorbidities, and COVID-19 PCR test results from nasal swab samples.[12] Additionally, the content of physician and physician assistant notes describing patients’ visits was reviewed.

### Measures

#### Outcome variable

The pilot model predicted acute COVID-19 infection at the individual patient level based on results of COVID-19 PCR testing performed during the ED visit.

#### Predictor variables

Selected predictor variables were covariates known to be associated with individual-level body temperature, including age, sex, month and time of day of temperature measurement, and comorbidities [8, 9, 13, 14]. Body temperature was operationally defined as the highest temperature measurement taken during the ED visit. Time of day of body temperature measurement ranged from 9:00AM to 9:00PM and was defined as a continuous variable, derived by squaring the nearest rounded hour of body temperature measurement. Comorbidities associated with individual variation in baseline body temperature and considered for inclusion as predictor variables were cancer, pulmonary disease, hypothyroidism, kidney disease, congestive heart failure, allergies, and depression [9, 13]. Comorbidities were enumerated through study team review of diagnoses listed in the patient’s past medical history.

#### Justification for using clinicians’ estimates of pretest probability without access to objective temperature data

Because body temperature is a key predictor for acute COVID-19 infection, we included it directly in our final model based on structured EHR data rather than asking clinicians to incorporate it in an overall pretest probability. To avoid using body temperature twice, we used clinicians’ estimates of pretest probability without access to objective body temperature data.

### Analyses

#### Derivation of Independent Variable

##### Physician pretest probability estimation

Two study team members trained in emergency medicine (CB and CL) independently reviewed clinical documentation (ED notes from physician assistants and physicians) to estimate pretest probability for COVID-19 while blinded to patients’ ED visit temperature measurements. Reviewers rated each case on a 5-point ordinal scale (1=20% suspicion or less; 5=80% suspicion or more). To determine estimates, reviewers were instructed to consider clinically relevant contextual factors, including local prevalence of COVID-19 at the time of the visit; patient-reported history of recent COVID-19 exposure within the last 14 days; patient-reported symptoms (e.g., fever, chills, cough, shortness of breath, chest tightness, headache, sore throat, fatigue, body aches, diarrhea, abdominal pain, confusion, loss of taste, loss of smell); and objective criteria (e.g., vital signs, excluding redacted temperature). In cases where reviewers’ estimates differed by exactly one point, the estimates were averaged to obtain a mean pretest probability. If estimates differed by more than one point, the two reviewers discussed the case until consensus was achieved. Physician pretest probability estimates were included as a continuous predictor variable in statistical modeling.

#### Statistical Analysis

Descriptive statistics were obtained using SPSS Statistics version 24.0 (IBM Corporation, Armonk, New York, USA) to compare demographics, comorbidities, and clinical characteristics of patients who tested positive and negative for acute COVID-19 infection. Between-group differences in demographics, comorbidities, and clinical characteristics were assessed using independent samples t-tests, Mann Whitney U tests, Pearson’s chi-square tests of independence, or Fisher’s exact tests. Intraclass correlation coefficients (ICCs) were calculated to estimate inter-rater agreement on physician pretest probability estimates [15, 16].

Bivariable and multivariable logistic regression models were fitted to the data to test the research hypotheses that independent variables are predictive of acute COVID-19 infection.

Three pilot models were estimated with the following combinations of predictor variables:

Model #1: Body temperature

Model #2: Clinical probability (Body temperature and pretest probability estimate)

Model #3: Enhanced clinical probability (Body temperature, pretest probability estimate, and body temperature-relevant covariates

Our primary hypothesis was that the predictive performance of Model #3 would exceed that of Model #2 due to the incorporation of body temperature-relevant covariates in Model #3. Predictive performance was evaluated using the area under the curve (AUC) and receiver operating curve (ROC) for discrimination, and calibration-in-large and calibration slope for calibration. Model performance, when evaluated in the same sample that was fitted, is overestimated. Therefore, we calculated the optimism of predictive performance measures using bootstrap to obtain optimism bias-corrected estimates.[17, 18] The significance of predictor main effects in bivariable and multivariable models were assessed using likelihood ratio chi-square tests, calculated as twice the difference of the log-likelihoods between the full model and the constrained model that that does not contain the effect. Differences in AUCs between models were tested for statistical significance using Delong’s Test with alpha set to 0.05 and using a one-sided test.[19] Regression analyses were carried out by the logistic procedure in SAS version 9.4 (SAS Institute Inc., Cary, North Carolina, USA). Calibration and optimism bias-corrected estimates were obtained using R version 4.0.3 (The R Foundation for Statistical Computing, Vienna, Austria). All study procedures were reviewed and approved by the Institutional Review Board at Cedars-Sinai Medical Center.

## RESULTS

Out of 128 patients evaluated for inclusion in the study, a total of 117 patients met inclusion criteria. (See Appendix Figure 1 for a study flow chart.) Forty out of 117 (34%) patients in the sample tested positive for COVID-19. Compared to patients who tested negative for COVID-19, patients who tested positive had higher maximum oral body temperatures in the emergency department (99.3 vs 98.5°F, p < 0.05), but demographic and comorbidity characteristics were generally similar to patients who tested negative (Table 1).

**Table 1.** Participant demographics

| *Characteristic* | *COVID-*<br>*(n=77)* | *COVID+*<br>*(n=40)* | *p-value* |
| --- | --- | --- | --- |
| Maximum BT from ED (°F), mean (SD)<sup>†</sup> | 98.5 (0.7) | 99.3 (1.3) | 0.002* |
| Pre-test probability estimate without BT,<br>median (range)<sup>‡</sup> | 3.5 (3.5) | 4.0 (2.5) | 0.01* |
| Age (years), mean (SD) | 43.2 (13.4) | 41.1 (14.7) | 0.442 |
| Female | 43 (55.8) | 19 (47.5) | 0.391 |
| English-speaking | 73 (94.8) | 36 (90.0) | 0.443 |
| Non-white | 17 (22.1) | 10 (25.0) | 0.722 |
| Hispanic | 11 (14.3) | 9 (22.5) | 0.425 |
| Number of comorbidities |  |  | 0.891 |
| 0 | 44 (57.1) | 21 (52.5) |  |
| 1 | 21 (27.3) | 12 (30.0) |  |
| ≥ 2 | 12 (15.6) | 7 (17.5) |  |
| Comorbidities<sup>**</sup> |  |  |  |
| Pulmonary disease | 7 (9.1) | 6 (15.0) | 0.353 |
| Cancer | 5 (6.5) | 6 (15.0) | 0.182 |
| Hypothyroidism | 6 (7.8) | 1 (2.5) | 0.420 |
| Depression | 4 (5.2) | 1 (2.5) | 0.659 |
| Kidney disease | 2 (2.6) | 1 (2.5) | 1.000 |
| Allergies | 3 (2.6) | 0 (0.0) | 0.550 |
| Congestive heart failure | 1 (1.3) | 0 (0.0) | 1.000 |

*Notes.* SD = standard deviation; BT = body temperature; ED = emergency department. Values reported as n (%) unless indicated.
\* $p \leq 0.05$
\*\*Two-sided fisher's exact test
<sup>†</sup>Welch's unequal variances t-test
<sup>‡</sup>Mann Whitney U Test

For the 117 patients in our study, COVID-19 pretest probability estimation by dual reviewers yielded identical scores in 55 (43.0%) cases, adjacent scores in 66 (51.6%) cases; and scores initially differing by more than 2 or more points in 7 (5.5%), which were resolved by adjudication. The intraclass correlation coefficient (ICC) for pretest probability estimation was 0.75 (95% CI 0.64-0.83).

After pretest probability estimates were determined, we used our data to derive three models of the relationship between body temperature and acute COVID-19 infection (Tables 2-4). Model #1, which used a patient’s maximum oral temperature alone, resulted in an AUC of 0.69 (95% CI 0.58-0.79). Model #2 used pretest probability estimates in addition to body temperature, and it achieved an AUC of 0.72 (95% CI 0.62-0.82, absolute difference in AUC between Model 2 versus 1 of 0.029 (95% CI −0.057-0.115, p=0.25)). Model #3, which approximated clinicians’ current method of developing a pretest probability by using the objective temperature data, pretest probability estimates, and body temperature-relevant covariates, yielded an AUC of 0.76 (95% CI 0.67-0.86, absolute difference in AUC between Model 3 versus 2 of 0.038 (95% CI −0.021 to 0.097, p=0.10), and optimism-adjusted AUC estimate of 0.672. See Table 2 for a list of AUCs for the three models, Table 3 for multivariable logistic regression coefficients and odds ratios predicting COVID-19 test result positivity under Model #3. See Figure 2 for a graphical representation of the ROC curves for Models #1-3.

**Table 2.** Models derived from pilot data (N = 117)

| *Model* | *Name* | *Data Source* | *AUC (95% CI)* |
| --- | --- | --- | --- |
| #1 | BT only | (structured data from EHR) | 0.69 (0.58 - 0.79) |
| #2 | Clinical probability | (#1 and pretest probability) | 0.72 (0.62 - 0.82) |
| #3 | Enhanced clinical probability | (#2 and BT-related covariates*) | 0.76 (0.67 - 0.86) |

*Notes.* AUC = area under the curve. BT = body temperature. CI = confidence interval.
\*Covariates included age, sex, month, time of day, and comorbidities. None had $p < 0.05$ .

**Table 3.** Multivariable logistic regression coefficients predicting COVID-19 test positivity under Model #3 (n=117)

| *Variable* | $\beta$ | *OR (95%CI)* | $SE_{\beta}$ | $\chi^2$ | *df* | *p* |
| --- | --- | --- | --- | --- | --- | --- |
| Maximum BT from ED (unit: °F) | 1.010 | 2.75 (1.43-5.25) | 0.331 | 9.325 | 1 | 0.002* |
| Pretest probability estimate<sup>†</sup> | 0.561 | 1.75 (1.01-3.05) | 0.282 | 3.975 | 1 | 0.046* |
| Age (unit: years) | 0.008 | 1.01 (0.98-1.04) | 0.017 | 0.263 | 1 | 0.608 |
| Sex (ref: Female) |  |  |  |  |  |  |
| Male | 0.669 | 1.95 (0.71-5.34) | 0.513 | 1.700 | 1 | 0.192 |
| Month of BT measurement (ref: March) |  |  |  |  |  |  |
| April | -0.397 | 0.67 (0.15-3.09) | 0.778 | 0.260 | 1 | 0.610 |
| Hour of BT measurement (unit: hour<sup>2</sup>) | -0.001 | 1.00 (0.99-1.01) | 0.003 | 0.074 | 1 | 0.786 |
| Comorbidities (ref: no disease) |  |  |  |  |  |  |
| Cancer | 1.450 | 4.26 (0.79-22.9) | 0.858 | 2.856 | 1 | 0.091 |
| Depression | 0.038 | 1.04 (0.10-11.4) | 1.221 | 0.001 | 1 | 0.975 |
| Hypothyroidism | -1.265 | 0.28 (0.02-3.46) | 1.279 | 0.978 | 1 | 0.323 |
| Kidney disease | -3.407 | 0.03 (0.00-5.46) | 2.604 | 1.711 | 1 | 0.191 |
| Pulmonary disease | 1.181 | 3.26 (0.77-13.7) | 0.734 | 2.587 | 1 | 0.108 |

*Notes.* $\beta$ = regression coefficient; BT = body temperature; df = degrees of freedom; ED = emergency department; OR = odds ratio; ref = reference category; SE = standard error. Chi-square statistics are Wald based.
\* $p \leq 0.05$
<sup>†</sup> Average of two raters' scores on a 5-point ordinal scale

**Figure 2.**
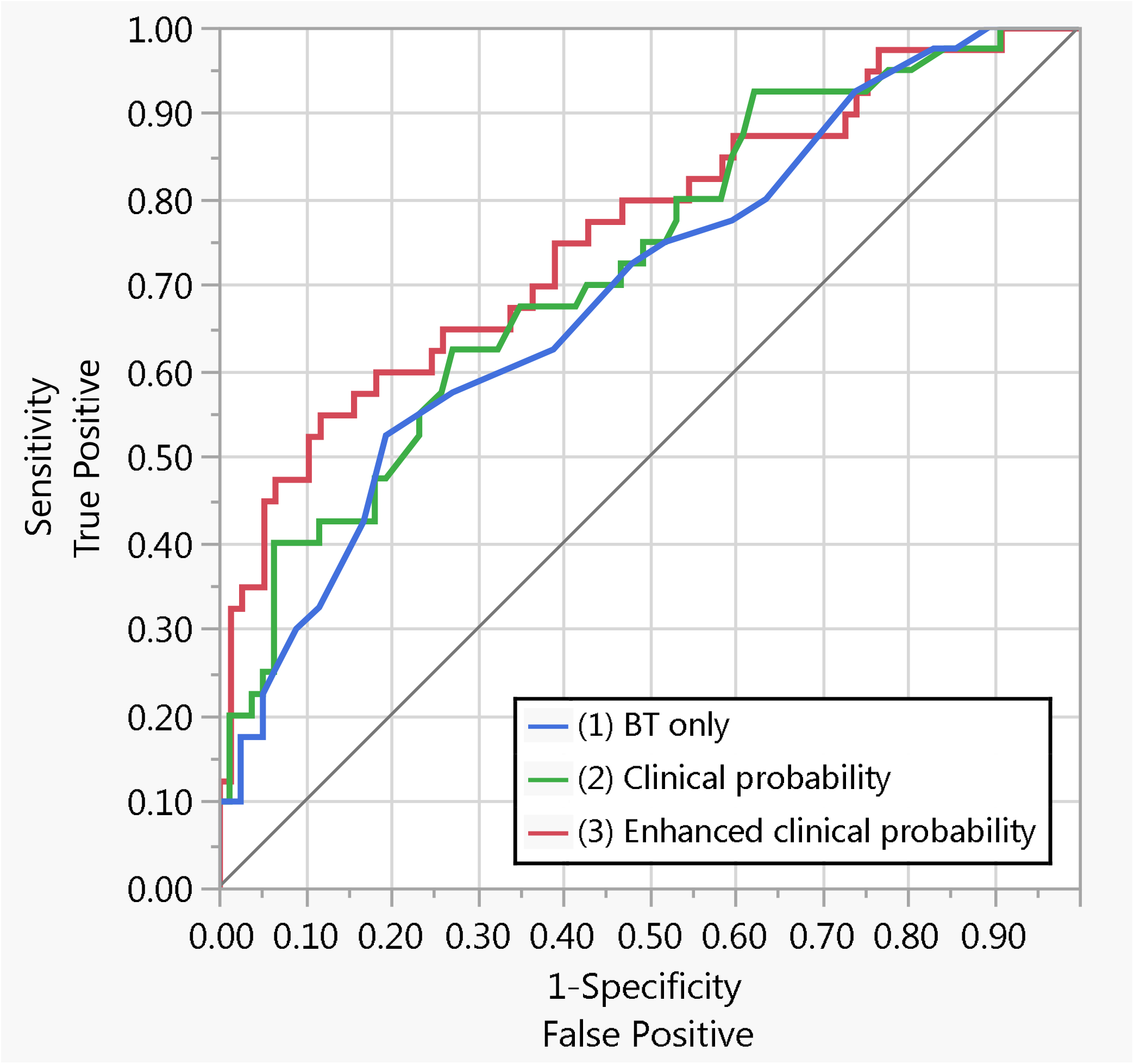
Receiver operating curves for Models #1-3

Apparent calibration slope and calibration-in-large for Model #3 were 1.0 and 0.0, respectively, indicating perfect calibration and overfitting. Bootstrap validation of Model #3 yielded an optimism-adjusted calibration slope of 0.478, indicating that the model underestimates low risk of COVID-19 infection and overestimates high risk of COVID-19 infection. Optimism-adjusted calibration-in-large was −0.293, indicating that the model on average overestimates the risk of a COVID-19 infection. See Figure 3 for calibration plots of apparent and optimism-adjusted Model #3.

**Figure 3.**
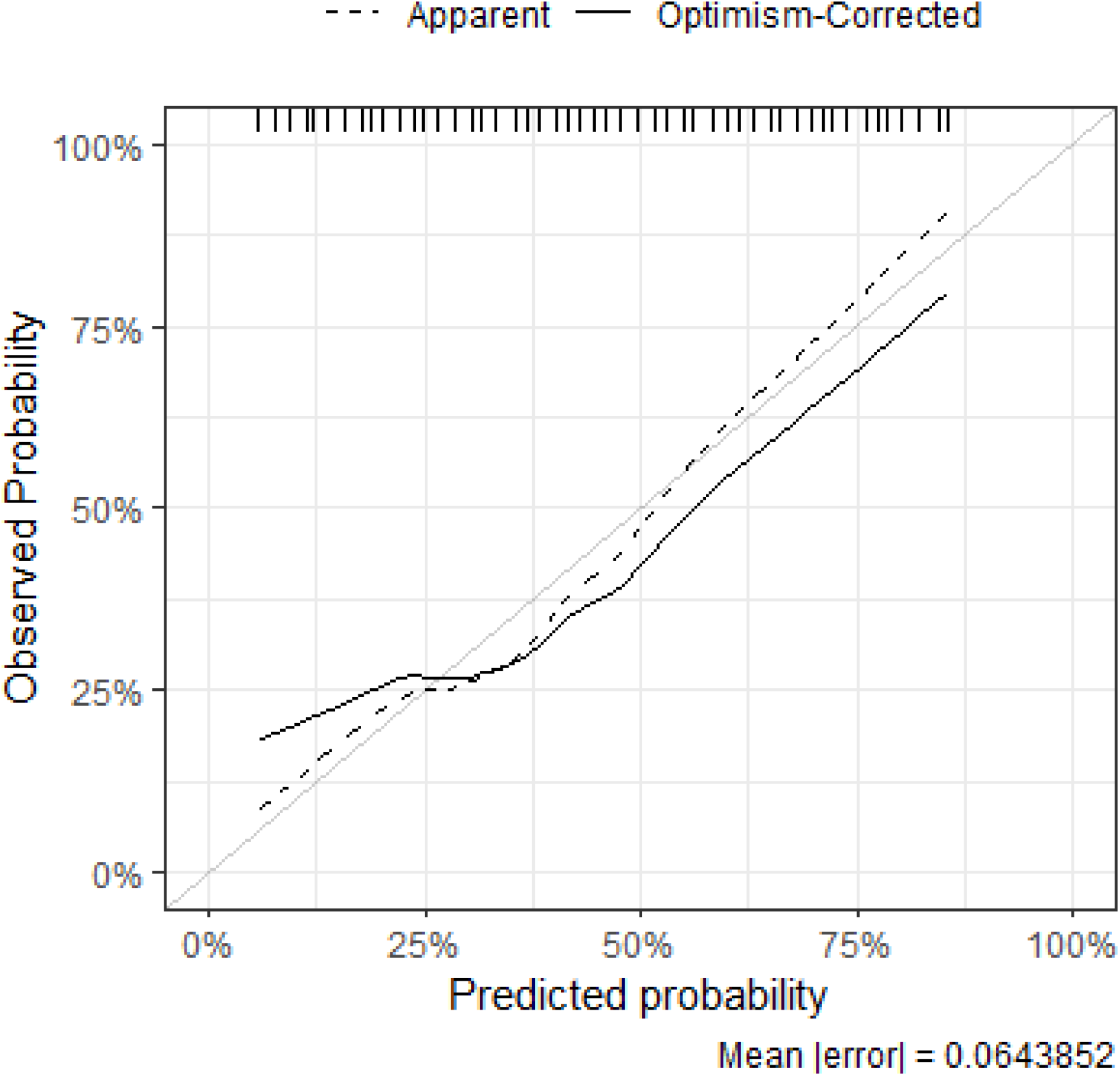
Calibration plot for apparent and optimism-adjusted Model #3

## DISCUSSION

Based on our *a priori* hypothesis that several covariates known to be associated with body temperature in non-infected subjects [8, 9, 13, 14] could improve the performance of body temperature as a predictor of COVID-19 acute infection status, we compared the predictive performance of three models. These models included oral body temperature alone, physicians’ temperature-blinded pretest probability plus objective temperature data, and a proposed enhanced model (which added age, sex, month, time of day, and comorbidities), which increased the AUC from 0.69 to 0.76—a statistically nonsignificant difference but a trend which should undergo investigation in further research.

Clinicians have long known that covariates are relevant to the interpretation of body temperature. Until now, however, most consideration of covariates’ effects has been handled qualitatively in clinicians’ minds. To the extent that these associations hold true and can be validated in other datasets, they provide a method for clinicians to quantitatively adjust their interpretation of body temperature which could be used to produce an evidence-based adjusted body temperature for every patient encounter. This could be likened to using a corrected QT interval, which has superseded use of an unadjusted QT interval in nearly all clinical settings. Because there are billions of patient encounters per year that use body temperature, the public health implications of even a tiny improvement in discriminatory ability are large.

We recognize that this initial analysis has several limitations. First, we expect that these AUCs are optimistic because they are evaluated in a derivation sample at a single site,[20] though we attempted to mitigate this effect by reporting optimism-adjusted performance measures of calibration and discrimination. Second, our data used a subset of patients who were tested for COVID-19 during a time when testing supplies were limited, making selection bias a potential limitation. Third, members of the research team who generated pretest probability estimates were not blinded to the study hypothesis regarding which covariates should affect interpretation of temperature. This may have caused their estimates to differ from estimates that would otherwise by generated in practicing clinicians. Fourth, even when AUC is increased, it must be considered in a clinical context to ensure that yields clinically significant benefit. Fifth, we were unable to include two important classes of variables in our analysis because of gaps in the sample: patients’ baseline temperature values and variables related to women’s hormonal cycles. Accounting for patients’ baseline temperature has been endorsed by the Infectious Disease Society of America[21]. Moreover, use of hormonal contraceptives and estrogen replacement therapy have been demonstrated to impact temperature interpretation,[7] which suggests that time since a woman’s last menstrual period may also affect temperature elevation in the setting of acute infection. These variables were infeasible for us to collect in our proof-of-concept study, but we believe that they would likely further enhance predictive performance of our model if we had been able to include them.

In conclusion, we present preliminary evidence that enhancing the interpretation of body temperature with body temperature-relevant covariates can improve discriminatory ability for COVID-19 above and beyond what is currently possible by using clinical suspicion and body temperature alone. If future research can validate these findings in other datasets, use of body-temperature relevant covariates to generate an adjusted temperature could be a promising method to improve detection of COVID-19 and other febrile illnesses, and to curb their spread.

## Data Availability

Data will be made available upon request, and only after permission from the Cedars-Sinai Medical Center Institutional Review Board.

## APPENDICES

**Appendix Figure 1.**
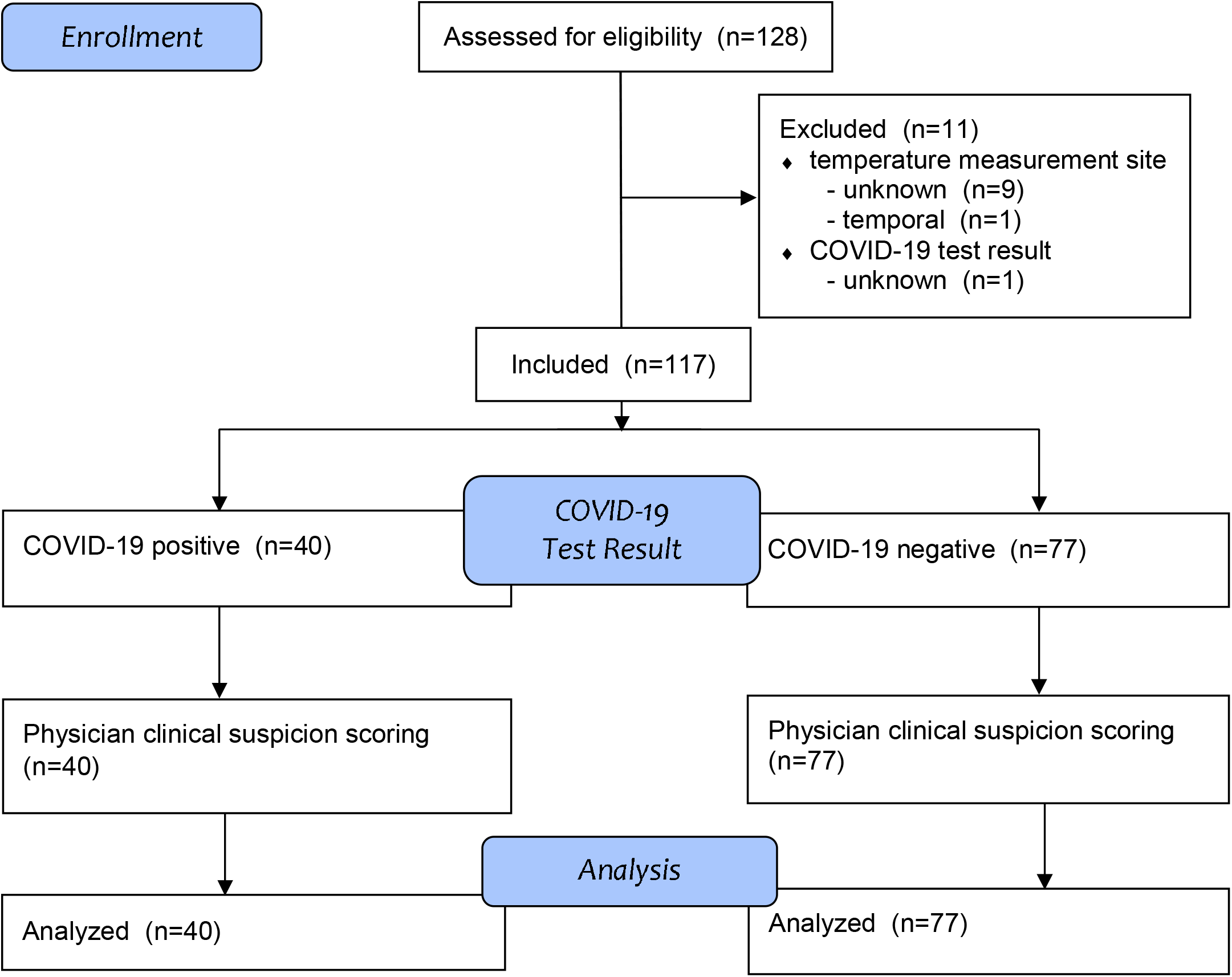
Study flow diagram

